# Levodopa Administration Timing During Hospitalization: Associations With Intensive Care Unit Exposure and Documented Access Type

**DOI:** 10.64898/2026.08.17.26360596

**Authors:** Alon Gorenshtein, Mark Katson, Yosef Adiniaev, Eyal Klang, Oved Daniel

## Abstract

**Background:** Levodopa is time-critical in hospitalized Parkinson disease. Whether dosing fidelity depends on care setting or documented access status is unclear.

**Objectives:** To quantify levodopa dosing fidelity, test ICU exposure with clustering-aware methods, and test whether documented access type is associated with delayed or omitted dosing.

**Methods:** Retrospective cohort study using MIMIC-IV (2011-2022). Adults with Parkinson disease and ≥1 scheduled levodopa dose contributed 1,665 admissions and 39,322 doses. ICU exposure was tested with a patient-clustered GEE model. Among ICU-exposed doses, access type (normal, tube feeding, parenteral nutrition, NPO) was modeled in one fully adjusted model and tested for specificity, restricted to the ICU, against an active-comparator medication (statins).

**Results:** Of 39,322 doses, 79.8% were on time by the primary 60-minute definition; a symmetric ±15-minute definition classified 68.8% as mistimed. ICU exposure was not associated with delayed or omitted dosing after clustering (patient-clustered OR, 0.87; 95% CI, 0.74-1.01). Among ICU-exposed doses, NPO was associated with delayed or omitted dosing (adjusted OR, 1.89; 95% CI, 1.36-2.62) and tube feeding with lower odds (adjusted OR, 0.62; 95% CI, 0.42-0.92; P < .001). The comparator medication showed a directionally consistent but inconclusive interaction (OR, 1.27-1.28; 92 patients). A route-order association was not observed among immediate-release formulations (OR, 0.72; 4 patients).

**Conclusions:** ICU admission alone was not associated with dosing unreliability after clustering. Among ICU-exposed doses, access type, not a single pooled category, was associated with dosing reliability; a comparator-medication check, valid only in the ICU, was directionally consistent but inconclusive.

## Introduction

Parkinson disease affects a growing number of adults worldwide, and its global prevalence more than doubled between 1990 and 2016.[1] Levodopa remains the most effective treatment for its motor symptoms, but its benefit depends on a narrow, patient-specific dosing schedule: a delayed or omitted dose can allow motor symptoms and rigidity to return within hours, and abrupt discontinuation risks parkinsonism-hyperpyrexia syndrome, a rare but life-threatening reaction distinct from dopamine-agonist withdrawal.[2] The Parkinson’s Foundation and allied guidance now recommend administering scheduled Parkinson disease medications within 15 minutes, stricter than the 60-minute cutoff used in most prior audits.[3] Levodopa is therefore a time-critical medication, and hospitals are increasingly encouraged to treat its scheduled administration as a safety priority.

Audits at individual hospitals have repeatedly found that scheduled Parkinson disease medications are delayed or omitted in a substantial minority of administrations, with reported rates of 10% to 30%.[4–6] These audits disagree on the downstream consequence: one reported a longer hospital stay among patients with a missed dosage,[4] another found no association.[5] Both were manual chart-review audits of a few hundred patients, lacking the scale and clustering-aware statistics an Electronic Medication Administration Record (EMAR) affords. Critically ill patients, who commonly have altered diet orders, ventilation, or sedative infusions that could interrupt oral levodopa delivery, have not been examined as a distinct population at this scale.[7] A recent Medical Information Mart for Intensive Care IV (MIMIC-IV) analysis examined mortality covariates among critically ill Parkinson disease patients but not administration timing.[8]

We conducted a retrospective cohort study using granular EMAR data to quantify dose-level levodopa fidelity, test the intensive care unit (ICU)-versus-ward association with clustering-aware methods, and test whether documented access type is associated with delayed or omitted dosing among ICU-exposed doses, with an active-comparator medication as a specificity check.

## Methods

### Study Design and Setting

This was a retrospective cohort study of adult hospital admissions with Parkinson disease at one US academic medical center, using MIMIC-IV, version 3.1, a de-identified electronic health record database.[9] The analytic window was restricted to the 2011-2022 anchor-year-group eras; the earliest era (2008-2010) was excluded after a preliminary comparison found it accounted for 69.0% of admissions with a diagnosis and levodopa order lacking any scheduled EMAR entry, versus 22.4% with one, consistent with incomplete EMAR rollout (eTable 1). Reporting followed STROBE for observational studies (eMethods; item map in eTable 2).[10] The quantity of interest was whether, when, and under what conditions scheduled levodopa doses were given as ordered; no causal effect on a clinical outcome was estimated.

### Cohort Identification

Admissions were included if they carried an ICD-9 or ICD-10 diagnosis code for idiopathic Parkinson disease (332.0 or G20) and had ≥1 carbidopa-levodopa order in the same admission, a single-code definition informed by an ascertainment approach previously validated in the Veterans Health Administration record, not in MIMIC-IV;[11] a stricter sensitivity cohort, requiring the code on ≥2 admissions, was also constructed. Every distinct matched drug string and formulary code was reviewed against its formulary description to confirm a genuine combination product (eTable 3). Admissions with only a secondary or drug-induced parkinsonism code (ICD-10 G21.x, ICD-9 332.1) were excluded, as were admissions whose levodopa order was never scheduled.

### The Dosing-Fidelity Measure

The unit of analysis was the scheduled dose, one EMAR record for a levodopa-containing medication, nested within admission and patient, classified using the EMAR event code and, where available, the scheduled-to-actual administration interval (eMethods, eTable 4).

“Administered” doses were delayed by at least 60 minutes or omitted (the primary definition) if the interval was 60 minutes or more, and on time otherwise; a negative interval (early) was rolled into on time for the primary definition, because the clinical concern here is late or omitted, not early, dosing. An unrecognized or blank event code (0.2%) was excluded. The classification was repeated at 15-, 30-, and 90-minute thresholds (15 matches current Parkinson’s Foundation guidance[3]); because the primary definition folds every early dose into on time, a symmetric mistimed definition (delayed, omitted, or early by at least the same window) was also constructed at 60 and 15 minutes.

ICU exposure was defined at the dose level: a scheduled dose was ICU-exposed if its scheduled time fell within any ICU stay for that admission, and ward-exposed otherwise, so one admission could contribute doses to both settings.

### Access-Type Classification

Among ICU-exposed doses, the most recent chartevents diet-type entry (itemid 224001) within a 12-hour lookback was classified into 4 ordered categories, reflecting whether an alternate route was already structurally in place: normal or texture-modified diet, tube feeding (a tube already sited), total or peripheral parenteral nutrition (TPN/PPN; no tube established), or NPO (the most restrictive, least-specified category) (eMethods). Because pooling all 3 restricted categories can combine opposite associations with fidelity (Results), the narrower NPO-only flag was the primary exposure; the pooled definition from earlier work (any of the 3, altered oral access) was a secondary comparison. The 12-hour lookback was tested against 6- and 24-hour alternatives (eTable 5).

Diet-type documentation is charted only in the ICU module (chartevents); no admission without an ICU stay contributed a single entry (0 of 1,340 zero-ICU admissions; eTable 6). Absence was therefore classified with normal access by construction, a reasonable default within an ICU stay but one that cannot distinguish a true normal diet from an undocumented one outside it; any comparison spanning ICU and ward doses is not interpretable as an access-type test (Comparator Medication Specificity Analysis).

Mechanical ventilation (procedureevents itemid 225792) and active sedative or analgesic infusion (propofol, midazolam, dexmedetomidine, or fentanyl) were flagged at the dose level and entered as covariates, with access category and era, in one prespecified model (Statistical Analysis). A discharge-summary text-concordance check for this variable is reported in the supplement (eMethods; eTable 20).

### Comparator Medication Specificity Analysis

To probe whether an access-type association was specific to levodopa, an active-comparator medication was used: administration timing of a second, routine drug class under the same access-type classification.[12] This design cannot rule out confounding as cleanly as a true negative control, because altered access could plausibly affect the comparator’s timing too; it instead asks whether the levodopa association is materially larger. Statins (atorvastatin, simvastatin) were selected a priori as the most common standing, non-PRN, oral medication class here with no time-critical window, extracted by the same EMAR pipeline as levodopa (eMethods). Because diet-type documentation is ICU-only (Access-Type Classification), only the ICU-restricted analysis is a valid specificity test; a full-shared-cohort version was also run for transparency but reported as descriptive only.

A GEE interaction model (medication x altered access, clustered by patient), restricted to ICU-exposed doses, tested whether the association differed between medications, with and without ventilation adjustment.

### Route Order and Formulation

The ordered route for each dose’s governing levodopa order was extracted from prescriptions.route and linked through the shared pharmacy identifier and formulary drug string; orders permitting enteral or sublingual administration were classified route-flexible, “PO” or “Oral“-only orders oral-only (eMethods). A formulation-by-route cross-tabulation showed every controlled-release (CR) dose in the altered-access subset was ordered oral-only; a CR tablet cannot be crushed for enteral administration, a pharmacological constraint, not a clinician choice. The association was re-estimated clustered by patient, restricted to the NPO-only subset and to immediate-release formulations to remove the CR confound, and cross-checked against the administered route in emar_detail for a random sample (eMethods).

### Landmark Outcomes and Ward-to-ICU Transfer

To reduce reverse causation, exploratory outcomes were assessed with a 24-hour landmark design, reported in the supplement (eMethods, eTable 7): dosing-fidelity burden in the first 24 hours was compared against length of stay, mortality, and a whole-admission ICD-coded composite across burden tertiles. The same structure, with exposure and outcome reversed, tested whether landmark-window burden was associated with a subsequent ICU transfer among admissions not yet ICU-exposed at the landmark (eMethods, eTable 8); this removes reverse causation from the timing of measurement but cannot exclude confounding by an unmeasured deterioration trajectory, so its result is hypothesis-generating only.

### Statistical Analysis

Dose-level fidelity proportions were summarized with 95% Wilson score confidence intervals. All GEE models used a binomial family, exchangeable working correlation, and clustering by patient. ICU exposure’s association was estimated with a covariate-free GEE model, reported as a patient-clustered OR and repeated under the symmetric mistimed definition; a within-admission comparison compared each admission’s ICU rate with its own ward rate (Wilcoxon signed-rank test). The primary access-type model used tube feeding, TPN/PPN, and NPO against normal access as reference, with ventilation, sedative infusion, and era as covariates in one prespecified model; a joint Wald test on the 3 coefficients’ cluster-robust covariance tested whether access type mattered as a set, and marginal probabilities are reported alongside the odds ratios (eTable 9). The pooled altered-access model added a single such indicator in place of the categorical term (aOR). Headline odds ratios were converted to approximate E-values.[13] The naive route-order comparison also used a chi-square test, Cramer V, and a Wald 95% CI; landmark length of stay used the Kruskal-Wallis test and epsilon-squared, mortality and the ICD-coded composite a chi-square test and Cramer V, with Benjamini-Hochberg adjustment. Primary-cohort proportions were re-estimated in the stricter ascertainment cohort. Significance threshold was P < .05, two-sided; analyses used Python 3.9 (pandas, NumPy, SciPy, statsmodels).

### Ethics

MIMIC-IV was approved by the institutional review boards of the Beth Israel Deaconess Medical Center and the Massachusetts Institute of Technology, with a waiver of individual consent because records are de-identified; access was through credentialed PhysioNet authorization. This secondary analysis did not require additional review board approval.

## Results

### Cohort

In the 2011-2022 anchor-year-group eras, 2,112 admissions among 1,138 patients met the primary cohort definition (Table 1; Figure 1). Of these, 1,665 admissions (78.8%) had ≥1 scheduled EMAR entry, contributing 39,322 doses among 934 patients; the remaining 447 (21.2%) had a levodopa order never scheduled, substantially lower than the unrestricted 40.2% (eTable 1). Of the 1,665 admissions, 290 (17.4%, 229 patients) had ≥1 ICU stay overlapping a scheduled dose; at the dose level, 6,201 of 39,322 doses (15.8%) were ICU-exposed.

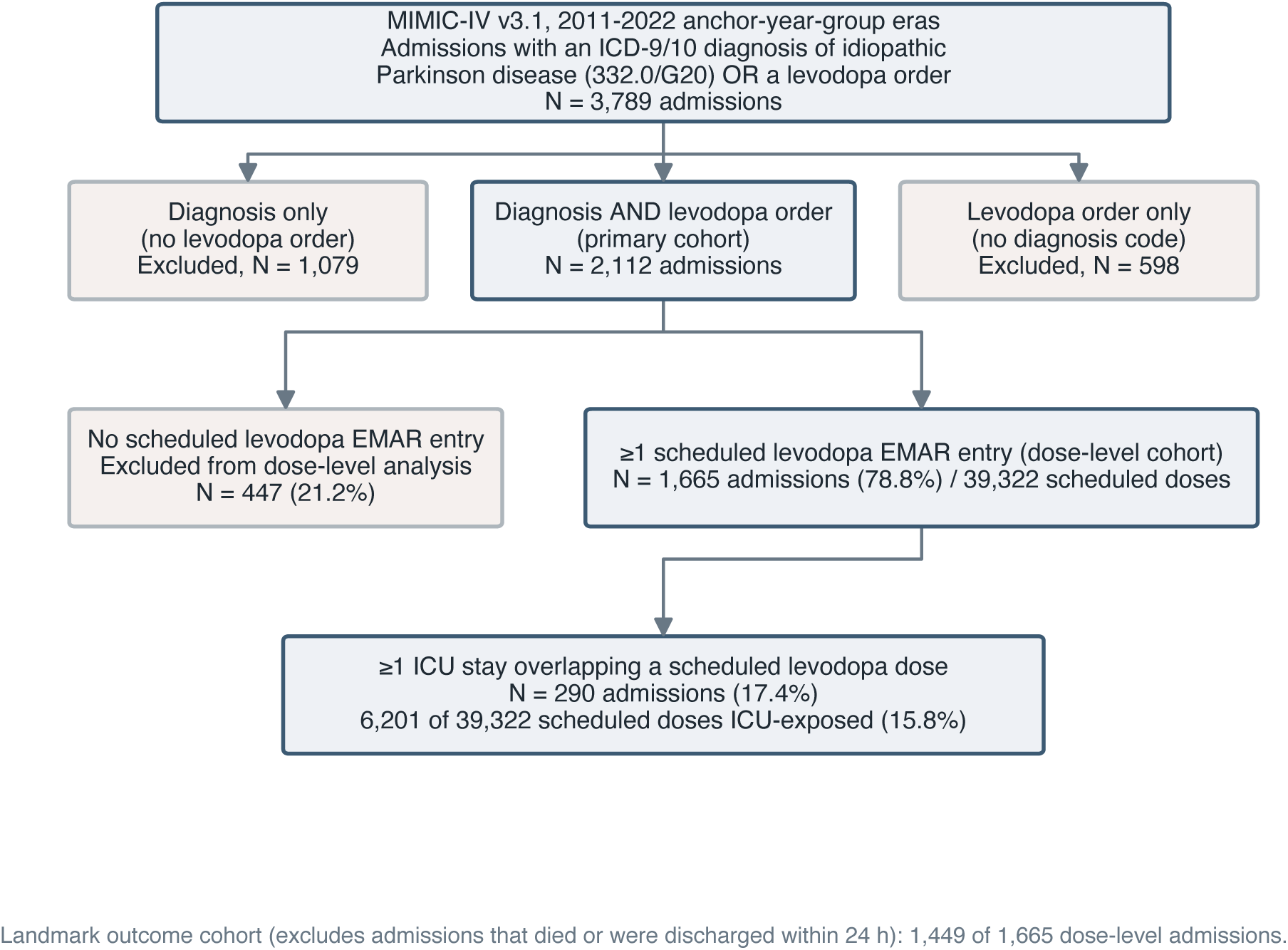

**Table 1.** Characteristics of the era-restricted Parkinson disease dosing-fidelity cohort.

| Characteristic | Value |
| --- | --- |
| Primary cohort (diagnosis + levodopa order, 2011-2022) |  |
| Patients, No. | 1,138 |
| Admissions, No. | 2,112 |
| Age, median (IQR), y | 76 (69-82) |
| Sex, No. (%) |  |
| Male | 720 (63.3) |
| Female | 418 (36.7) |
| Dose-level cohort ($\geq 1$ scheduled levodopa dose) | |
| Admissions, No. (%)<sup>a</sup> | 1,665 (78.8) |
| Admissions with no scheduled dose, No. (%)<sup>a</sup> | 447 (21.2) |
| Patients, No. | 934 |
| Scheduled levodopa doses, No. | 39,322 |
| Doses per admission, median (IQR) | 15 (8-28) |
| In-hospital mortality, No. (%) | 46 (2.8) |
| ICU-exposed subset<sup>b</sup> |  |
| Admissions with ICU exposure, No. (%) | 290 (17.4) |
| Patients with ICU-exposed doses, No. | 229 |
| Doses with ICU exposure, No. (%) | 6,201 (15.8) |
| With altered oral access, No. (%)<sup>c</sup> | 4,926 (79.4) |
| With mechanical ventilation, No. (%)<sup>c</sup> | 2,109 (34.0) |
| With active sedative infusion, No. (%)<sup>c</sup> | 624 (10.1) |
| Data-quality checks |  |
| Administered doses with no usable timestamp, No. (%) | 0 (0.0) |
| Doses coded "Partial Administered," No. (%) | 0 (0.0) |

<sup>a</sup>Among the 2,112 primary-cohort admissions. <sup>b</sup>Among the 1,665 admissions (39,322 doses) with a scheduled levodopa dose. <sup>c</sup>Among the 6,201 ICU-exposed doses. ICU indicates intensive care unit; IQR, interquartile range.

### Dosing Fidelity and Early Administration

Of 39,322 scheduled doses, 31,366 (79.8%; 95% CI, 79.4-80.2) were on time by the primary definition, 5,449 (13.9%) were delayed ≥60 minutes, and 2,507 (6.4%) were omitted, coded “Not Given” (2,412) or “Hold Dose” (95) (Figure 2); no administered dose lacked a usable timestamp and none was coded “Partial Administered” (eTable 4). Of the on-time doses, 11,263 (28.6% of all doses) were early, median magnitude 17 minutes (IQR, 8-32); ≥60 minutes early was rare (0.3%). Under the symmetric ±15-minute definition, 68.8% of doses were mistimed, materially more than the 20.2% mistimed by the primary 60-minute definition; the primary definition was retained for that reason (eTable 10). Early administration was modestly more common in the ICU than ward (32.4% vs 27.9%).

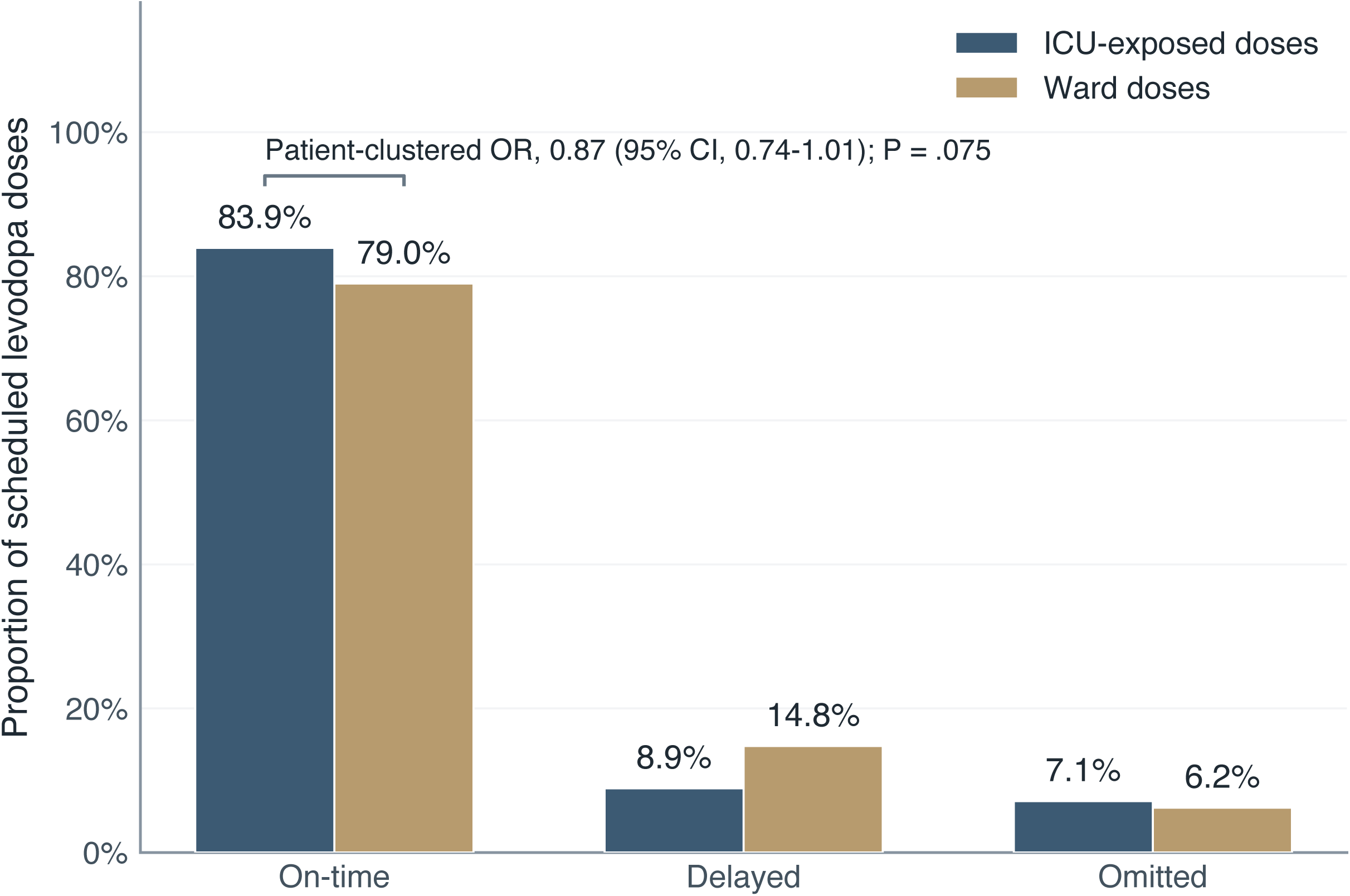

### Setting: ICU Exposure Was Not Associated With Dosing Fidelity Once Clustering Was Addressed

The unadjusted delayed-or-omitted rate was lower among ICU-exposed than ward doses (16.1% vs 21.0%). After clustering by patient, ICU exposure was not associated with delayed or omitted dosing (patient-clustered OR, 0.87; 95% CI, 0.74-1.01; P = .075); this CI does not establish equivalence (E-value, 1.08; eTable 11). Among 249 admissions with doses in both settings, the within-admission ICU rate was numerically lower but not significantly so (Wilcoxon, P = .11) (Figure 2). The unadjusted direction held at every delay threshold (eTable 12) and under the symmetric definition (60 minutes, OR 0.86; 15 minutes, OR 1.05, closer to null) (eTable 10).

### Access Type Among ICU-Exposed Doses

Among the 6,201 ICU-exposed doses, access type was normal or texture-modified in 1,275 (17.0% mistimed), tube feeding in 3,084 (9.6%), TPN/PPN in 35 (20.0%), and NPO in 1,807 (26.3%). In the fully adjusted, patient-clustered model, tube feeding was associated with lower odds of delayed or omitted dosing (aOR, 0.62; 95% CI, 0.42-0.92) and NPO with higher odds (aOR, 1.89; 95% CI, 1.36-2.62) than normal access (Figure 3; eTable 13); a joint

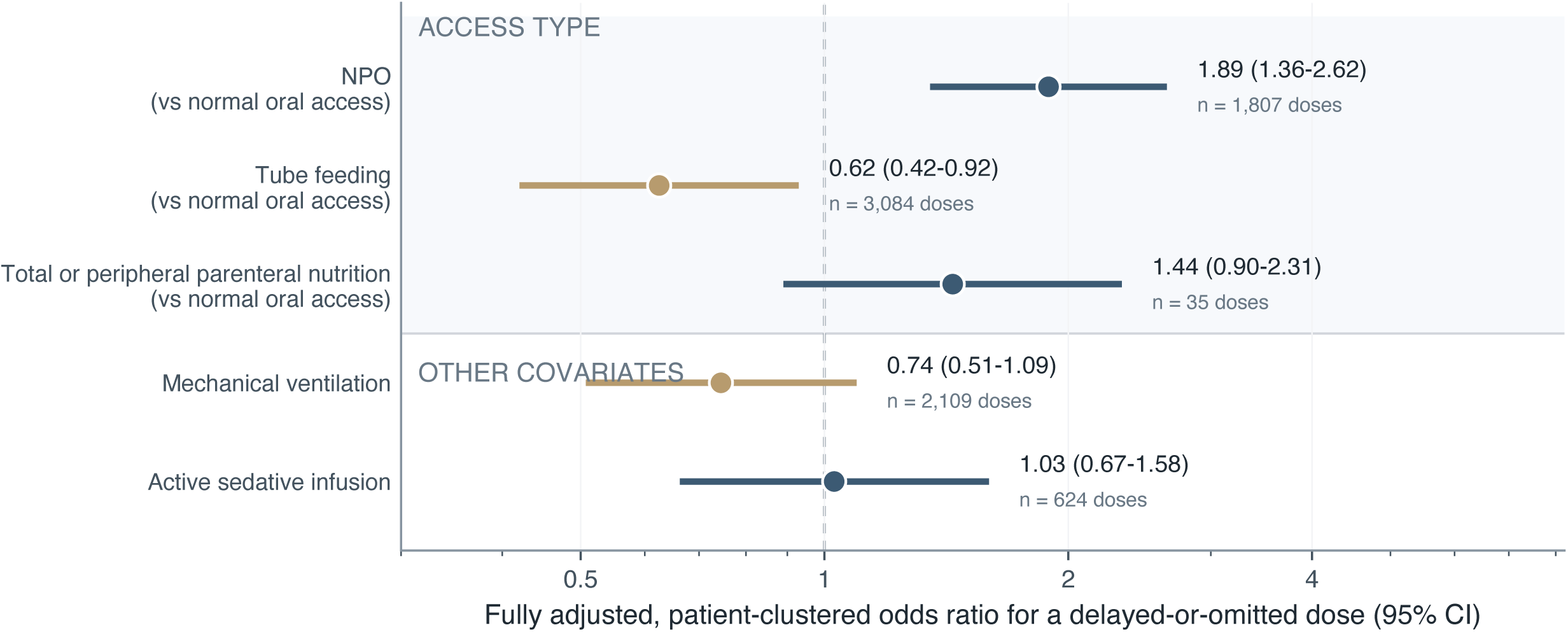

Wald test confirmed access type mattered as a set (χ^2^3 = 72.5; P < .001). TPN/PPN showed a similar but nonsignificant direction from only 35 doses and 2 patients, too few for inference (aOR, 1.44; 95% CI, 0.90-2.31) (eTable 13). Predicted probabilities of a delayed- or-omitted dose ranged from 10.1% (tube feeding) to 25.4% (NPO), against 15.3% for normal access (Figure 4; eTable 9). The narrower NPO-only definition, the primary exposure, showed a comparably strong unadjusted association (OR, 2.24; 95% CI, 1.88- 2.67), stable across a 6- or 24-hour lookback (eTable 5). The pooled altered-access definition attenuated this (aOR, 1.43; 95% CI, 1.09-1.87; E-value, 1.26) by folding the protective tube-feeding category with the harmful ones (eTable 14).

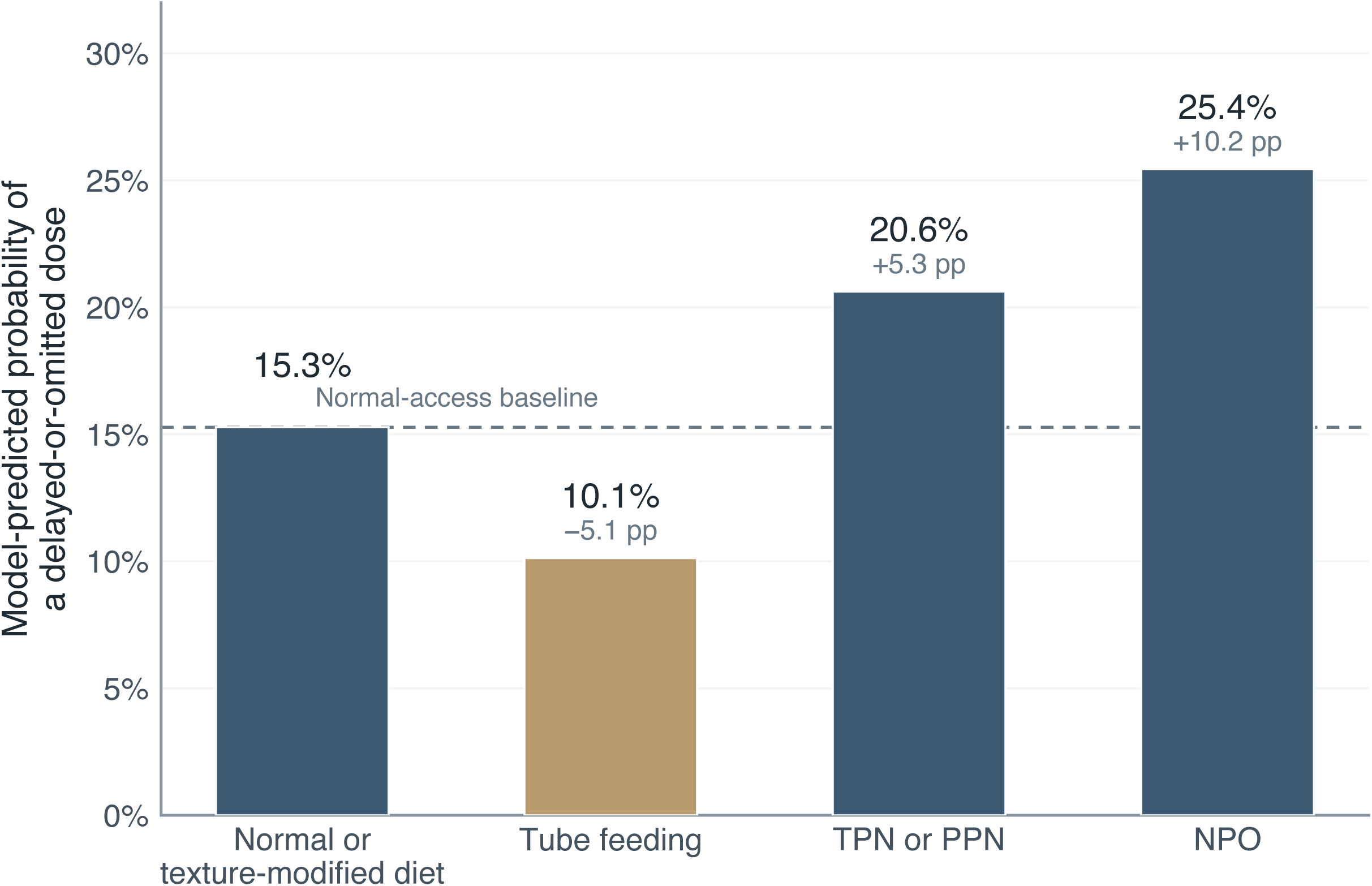

Mechanical ventilation was strongly associated with altered access (98.6% of ventilated doses vs 69.6% of non-ventilated; Cramer V, 0.34; P < .001); its own association attenuated to nonsignificance once access type was modeled as a category rather than pooled (aOR, 0.74; 95% CI, 0.51-1.09; P = .13) (eTable 13), against a significant, independently protective association in the earlier pooled model (aOR, 0.71; 95% CI, 0.56-0.90; eTable 15). Sedative infusion was not associated with mistiming in either specification (aOR, 1.03; 95% CI, 0.67-1.58). A discharge-summary text-concordance check corroborated normal access (88.3% specificity) and tube feeding (71.9% sensitivity) but rarely NPO (17.6% sensitivity) (eTable 20).

### Specificity: An Active-Comparator Medication, Restricted to the ICU

Only the comparison restricted to ICU-exposed doses is interpretable as a specificity test (Methods; eTable 6). In the 92 patients with both a scheduled levodopa and statin dose during ICU exposure, the medication-by-access interaction was directionally consistent with a levodopa-specific effect but not statistically significant, with or without ventilation adjustment (OR, 1.27-1.28; 95% CI crossing 1; P = .36-.37) (eTable 16). The same classification was also applied across the full shared cohort of 653 admissions (394 patients): both medications appeared more reliable under altered access, less so for levodopa, producing a significant interaction (OR, 1.66; 95% CI, 1.21-2.27; P = .002; 20,993 doses) (eTable 16). This full-cohort estimate is descriptive only, since ward-only admissions could never be flagged altered access regardless of true diet status.

### Route Order: An Exploratory Finding Confounded by Formulation

Among the 4,601 ICU-exposed doses with altered access and a classifiable route (180 patients), oral-only orders (71 doses) were delayed or omitted more often than route-flexible orders (4,530 doses; 22.5% vs 9.7%; OR, 2.72; 95% CI, 1.54-4.78; E-value, 1.79), attenuating after clustering (OR, 2.33; 95% CI, 1.27-4.26). A formulation-by-route cross-tabulation showed 39 of the 71 oral-only doses (54.9%) were controlled-release, versus none of the route-flexible doses; controlled-release tablets cannot be crushed for enteral administration, a pharmacological constraint, not a clinician choice. Restricted to immediate-release formulations, the point estimate reversed but only 4 patients contributed an oral-only dose (OR, 0.72; 95% CI, 0.30-1.73), too imprecise for a conclusion. A cross-validation against emar_detail’s administered route found 10.6% of sampled route-flexible doses were actually given non-orally (eTable 17; eTable 18).

### Landmark Outcomes, Ward-to-ICU Transfer, and Sensitivity Analyses

Landmark-restricted analyses, in the supplement, found a small length-of-stay association across dosing-fidelity burden tertiles (epsilon-squared, 0.007; BH-adjusted P = .006) and no association with mortality or a whole-admission ICD-coded composite (BH-adjusted P = .32 and .15) (eTable 7). Reversing exposure and outcome, landmark-window burden was not associated with a subsequent ICU transfer at 24 or 48 hours (OR, 0.91-1.29; 12-20 events) (eTable 8). Dose-level fidelity proportions were similar in the stricter PD-ascertainment cohort (73.1% of doses): 80.4% on time, 13.5% delayed, 6.1% omitted, versus 79.8%, 13.9%, 6.4% in the primary cohort (eTable 19), and unchanged restricting further to 2014-2022 (eTable 1).

## Discussion

In this cohort study, roughly one in five scheduled levodopa doses was delayed by at least 60 minutes or omitted, and more than two-thirds were mistimed under a stricter, symmetric 15-minute definition. Once patient clustering was addressed, ICU exposure was not associated with dosing unreliability. Among ICU-exposed doses, access type was the strongest correlate: tube feeding was associated with more reliable dosing than even normal access, while NPO was associated with less reliable dosing; the pooled altered-access definition from earlier work on this cohort attenuated this pattern by folding the protective and harmful categories together. An active-comparator medication, restricted to the ICU doses where the classification is valid, showed a directionally consistent but statistically inconclusive difference (OR, 1.27-1.28; 92 patients). An unadjusted route-order association could not be distinguished from formulation confounding and is best regarded as exploratory.

The route-order finding illustrates a related but distinct hazard: an order-level exposure that looks like a modifiable design choice can instead reflect a formulation constraint, since restricting to immediate-release formulations left too few oral-only patients for a conclusion.

Ventilation’s association with dosing fidelity attenuated to nonsignificance once access type was modeled directly rather than pooled, consistent with much of its earlier apparent independent effect reflecting its correlation with tube feeding rather than an independent mechanism; closer nursing or respiratory-therapy attention, an established access device, or unmeasured ICU staffing intensity could all still contribute. Reversing the landmark design found no evidence that dosing-fidelity burden predicted a subsequent ICU transfer, though with few events and residual confounding.

Prior single-center chart-review audits reported 10% to 30% of scheduled Parkinson disease medication doses delayed or omitted.[4–6] The proportion here (16.1%-21.0% by setting) falls within that range but was derived from EMAR timestamps across more than a decade with clustering-aware statistics; under a stricter, guideline-concordant definition, most doses were mistimed. Two prior audits disagreed about whether a missed dose lengthened hospital stay;[4,5] the landmark analysis found a substantially smaller effect and no association with mortality, the ICD-coded composite, or a subsequent ICU transfer, consistent with dosing fidelity tracking severity as much as, or instead of, changing outcomes. A recent MIMIC-IV analysis in a small critically ill cohort examined mortality covariates but not timing.[8]

These findings suggest pharmacy and nursing interventions might be more productively targeted at ensuring an alternate route is established when oral access is restricted, for example an alert when NPO status is newly documented with no enteral access in place, than at the ICU as a setting or route flexibility. This retrospective study cannot establish that such an alert would reduce delay or omission, only that access type and dosing reliability are associated here; the comparator-medication check, valid only within the ICU, was directionally consistent but too imprecise to confirm it.

### Limitations

This study has several limitations. First, this was a single-center study using one EHR and nursing workflow, so associations here may not generalize. Second, access type came from a semi-structured free-text field, not a validated flag or device-presence record: “NPO” does not prove a feeding tube was absent, and “tube feeding” does not prove the tube was usable for a given formulation; access type is best read as documented status, with route availability a plausible explanation, not a measured exposure. This study also had no clinician available to adjudicate held or delayed doses; an automated discharge-summary check corroborated the normal and tube-feeding categories but rarely NPO, consistent with NPO being transient or implied by other findings rather than narratively documented (eTable 20). Third, the comparator-medication analysis, restricted to ICU-exposed doses where documentation is valid, was underpowered (92 patients) and inconclusive; it used a single drug class chosen a priori and is an active-comparator check, not a true negative control. Fourth, the TPN/PPN category contained only 35 doses from 2 patients, reported descriptively. Fifth, the route-order analysis could not fully separate order-level from dose-level exposure; restricted to immediate-release formulations, only 4 oral-only patients remained. Sixth, MIMIC-IV has no validated severity score or outpatient timing data, so the landmark and ward-to-ICU comparisons remain vulnerable to unmeasured confounding. Seventh, the cohort was restricted to levodopa and idiopathic Parkinson disease.

### Future Directions

A multicenter replication large enough to analyze route flexibility at the order level within a single formulation class would test that association independent of the confound identified here. A structured, chart-validated access-type flag documented hospital-wide would allow a full-cohort specificity test and direct assessment of route availability rather than documented diet status. Prospective evaluation of an access-type alert, ideally stepped-wedge, would test whether a targeted workflow change reduces delayed or omitted dosing.

## Conclusion

In this cohort study, ICU admission alone was not associated with unreliable levodopa dosing after patient clustering. Among ICU-exposed doses, access type was associated with dosing reliability; a comparator-medication check restricted to the ICU, where the comparison is valid, was directionally consistent but statistically inconclusive. An apparent association with the ordered route could not be distinguished from formulation confounding and requires confirmation in a larger, order-level analysis.

## Data Availability

MIMIC-IV v3.1 is available to credentialed researchers through PhysioNet (https://physionet.org/content/mimiciv/) after completion of human-subjects training and execution of the data use agreement; it cannot be redistributed. Analysis code (cohort extraction, dose-level and admission-level dataset construction, route and access classification, the access-type categorical and lookback-sensitivity analysis, the comparator-medication specificity analysis, the route-order re-analysis and formulation-confound check, landmark and ward-to-ICU-transfer dataset construction, E-value computation, statistical analysis, and figure generation) is available at https://github.com/Alon-Gorenshtein/Levodopa-Administration-Timing-During-Hospitalization. The repository does not contain patient-level data, consistent with the MIMIC-IV data use agreement.

https://github.com/Alon-Gorenshtein/Levodopa-Administration-Timing-During-Hospitalization

## Funding

AG and EK were supported in part by the Clinical and Translational Science Awards (CTSA) grant UL1TR002541 from the National Center for Advancing Translational Sciences, through the Harvard Catalyst | The Harvard Clinical and Translational Science Center Pilot Award Program. The content is solely the responsibility of the authors and does not necessarily represent the official views of the National Institutes of Health.

## Competing interests

The authors declare that they have no competing interests.

## Notes

### Competing Interest Statement

The authors have declared no competing interest.

