## Supplementary information for "Levodopa Administration Timing During Hospitalization: Associations With Intensive Care Unit Exposure and Documented Access Type"

This Supplementary Information accompanies the main manuscript. It contains the STROBE item map, supplementary methods detail on cohort construction, era restriction, covariate extraction, the access-type categorical classification and lookback sensitivity, the comparator-medication specificity analysis, the route-order re-analysis and formulation-confound check, timing-sensitivity analyses, the landmark and ward-to-ICU-transfer design, and a discharge-summary text-concordance check for the access-type variable; supplementary tables reporting the excluded-admission comparison, the fine-grained dose-classification breakdown, the access-type categorical model, the comparator-medication analysis, E-values for the headline estimates, the route-order evidence ladder and cross-validation, landmark, ward-to-ICU, and ascertainment sensitivity results, and the text-concordance check; a codebook of the ICD codes, drug-name matches, and MIMIC-IV itemids used to build the cohort; and the code and data availability statement.

Contents

S1 Supplementary Methods S2 Supplementary Tables (eTable 1 to eTable 20) S3 Codebook S4 Code and Data Availability

### S1 Supplementary Methods

#### S1.1 STROBE Reporting

Reporting followed the STROBE statement for observational cohort studies. eTable 2 maps each STROBE item to its location in the main manuscript or this Supplementary Information.

#### S1.2 Cohort Construction and Era Restriction Detail

Admissions were identified from MIMIC-IV v3.1 hosp-module tables. Idiopathic Parkinson disease diagnoses were identified from diagnoses_icd using ICD-9 code 332.0 and ICD-10 code G20; secondary or drug-induced parkinsonism codes (ICD-10 G21.x, ICD-9 332.1) were not included. Levodopa orders were identified from prescriptions by a case-insensitive match of the drug field to "carbidopa" (eTable 3 lists the specific formulary strings matched). Every matched string, including the generic-labeled "carbidopa (generic entries)" row, was individually reviewed against its formulary description in prescriptions.formulary_drug_cd and prescriptions.prod_strength before inclusion; each confirmed a genuine carbidopa-levodopa combination product (immediate-release or controlled-release), not a carbidopa-only or unrelated product. The primary cohort required both a qualifying diagnosis code and a qualifying prescription in the same admission (hadm_id).

Before era restriction, 3,589 admissions met this definition, of which 1,444 (40.2%) had no scheduled electronic medication administration record (EMAR) entry for levodopa. eTable 1a compares included and excluded-no-EMAR admissions by anchor_year_group: the 2008-2010 era accounted for 69.0% of excluded-no-EMAR admissions but only 22.4% of included admissions, a pattern consistent with incomplete EMAR rollout in the earliest era rather than uniform order discontinuation across eras. Age, sex, and in-hospital mortality were similar between the two groups. Based on this finding, the analytic window was restricted to the 2011-2022 anchor-year-group eras for all primary analyses; within this restricted window, 2,112 admissions met the primary definition, of which 1,665 (78.8%) had a scheduled EMAR entry, a substantially smaller gap than the unrestricted 40.2%. A residual, smaller imbalance remained in 2011-2013; eTable 1b reports a further sensitivity restricting the primary cohort to 2014-2022 only, which produced materially unchanged proportions. The remaining gap could not be validated against a chart-level ground truth in MIMIC-IV and is noted as a limitation in the main manuscript.

#### S1.3 Dose-Level Fidelity Classification

Levodopa administration records were extracted from emar by matching the medication field to "levodopa" (case-insensitive), restricted to the era-restricted cohort's admission identifiers. Each record's event_txt field and its scheduletime and charttime fields were classified into a fine-grained category (on_time, early, delayed, omitted_not_given, omitted_hold, partial, or missing_timestamp_administered) before being rolled up to the three-way primary category used in the main text (eTable 4). Of 39,418 extracted records in the era-restricted cohort, 96 (0.2%) carried an unrecognized or blank event_txt value and were dropped before analysis. No record was coded "Partial Administered," and no "Administered"-coded record lacked a usable timestamp pair; both counts are exact zeros, not rounded small numbers.

#### S1.4 Critical-Illness Covariate Extraction

ICU exposure intervals came from icustays (intime to outtime). Mechanical ventilation intervals came from procedureevents restricted to itemid 225792 (invasive ventilation). Sedative and analgesic infusion intervals came from inputevents restricted to itemids 222168 (propofol), 221668 (midazolam), 225150 and 229420 (dexmedetomidine), and 221744, 225942, and 225972 (fentanyl).

Admission era used each patient's MIMIC-IV anchor_year_group field (five levels: 2008-2010, 2011-2013, 2014-2016, 2017-2019, 2020-2022); the earliest level was excluded from the analytic cohort (S1.2). Raw admittime values are shifted by a random, patient-specific offset and are not comparable across patients as calendar time, so anchor_year_group was used for any secular-trend adjustment instead.

#### S1.5 Access-Type Categorical Classification and Lookback Sensitivity

Altered oral access came from chartevents itemid 224001 ("Diet Type", a free-text field), which is charted exclusively by the ICU nursing system: no admission without an ICU stay contributed a single diet-type entry (0 of 1,340 zero-ICU admissions versus 322 of 325, 99.1%, of ICU-exposed admissions; eTable 6). Absence of an entry was classified with normal access by construction throughout, since a diet order not yet charted within an ICU stay is usually a genuinely unrestricted diet; outside the ICU, however, this means "normal access" reflects the absence of the source table rather than a documented normal diet, which is why any comparison spanning ICU and ward doses (S1.6) is reported as descriptive only, not as a specificity test.

Round 2/3 of this study used a single flag, active if the most recent diet-type entry within a 12-hour lookback contained "NPO", "Tube Feeding", "TPN", or "PPN" (case-insensitive), termed altered oral access (widened definition); a narrower definition restricted to the "NPO" substring alone was also computed (eTable 14). This round replaced the pooled flag as the primary exposure with a 4-level ordered categorical classification, reflecting whether an alternate administration route was already structurally in place at the time of the dose rather than nutritional severity: level 0, normal or any texture-modified diet value (Pureed, Clear liquid, Nectar/Honey Thick, Soft Solid, Thin Liquid, Full Liquid, Sips, Ground, or no diet-type entry in the lookback); level 1, tube feeding (an enteral tube already sited, so a route-flexible medication order can use it without a new access procedure); level 2, total or peripheral parenteral nutrition (TPN/PPN; no tube established); level 3, nil per os (NPO), the most restrictive and least-specified category, with no established alternate route. This ordering was pre-specified by structural rationale, not derived from the outcome data; eTable 13 reports that the resulting association with dosing fidelity was not monotonic in this ordering (tube feeding was more reliable than normal access), so the categorical model, not a forced linear trend, is the primary access-type analysis. A joint Wald test on the 3 access-category coefficients' cluster-robust covariance, from the fully adjusted model (S1.8), tested whether access type mattered as a set (eTable 13c); this replaced a round-4 omnibus Kruskal-Wallis test that had been run on raw dose-level values without accounting for patient clustering, the same pseudoreplication problem this round's statistics were designed to avoid elsewhere.

Both the categorical and the narrow/widened binary definitions used a 12-hour lookback window as the primary choice. As a sensitivity analysis, the narrow (NPO-only) flag was re-derived at 6- and 24-hour lookback windows; at each window, the fraction of ICU-exposed doses with no diet-type documentation at all in the lookback (as opposed to a documented normal-diet entry) was also computed (eTable 5). The 12-hour window was retained as primary because it had substantially better documentation coverage (96.8% of ICU-exposed doses had some diet-type entry in the lookback) than the 6-hour window (80.1%), while the association's magnitude was materially unchanged across all 3 windows tested.

#### S1.6 Comparator Medication Specificity Analysis

An active-comparator medication was used to probe whether the access-type association with dosing fidelity was specific to levodopa; because altered oral access could plausibly affect the comparator's own timing too, this design cannot rule out confounding as cleanly as a true negative control, and instead asks whether the levodopa association is materially larger than the comparator's. Statins were selected a priori as the comparator class: almost always standing, non-PRN, once-daily oral orders in this cohort (avoiding a scheduled-versus-PRN confound that a PRN-heavy comparator, such as PRN acetaminophen, would carry), with no accepted time-critical administration window, and among the most common non-fluid, non-electrolyte medication classes co-prescribed to this cohort after levodopa itself. Atorvastatin and simvastatin EMAR records were extracted by the same pipeline used for levodopa (event_txt and scheduletime/charttime classification), restricted to the subset of the levodopa cohort's admissions that also had ≥1 scheduled statin dose (653 of 1,665 admissions). Altered access (widened binary definition) and ICU exposure were joined to comparator doses identically to levodopa. A GEE logistic model with terms for medication (levodopa versus comparator), altered access, and their interaction, clustered by patient, tested whether the altered-access association with mistiming (delayed-or-omitted, 60-minute definition) differed between the two medications, restricted to ICU-exposed doses, with and without a mechanical-ventilation covariate; because diet-type documentation is ICU-only (S1.5), only this ICU-restricted version is a valid specificity test. The same model was also fit on the full shared cohort (all doses, ICU and ward) for transparency, but that estimate is reported as descriptive only, since ward-only admissions can never be classified altered access regardless of true diet status (eTable 16).

#### S1.7 Route-Order Extraction and Formulation-Confound Re-Analysis

The ordered administration route came from prescriptions.route, linked to each emar record through the shared pharmacy_id (emar.pharmacy_id to prescriptions.pharmacy_id); the formulary drug string for the same pharmacy_id was extracted alongside it. Route strings "PO/NG", "NG", "G TUBE", "J TUBE", and "SL" were classified route-flexible; "PO" and "ORAL" were classified oral-only; any other or missing route string was classified unknown and excluded from the route-order analysis.

This re-analysis: (1) reported the number of unique patients, admissions, and distinct pharmacy orders (pharmacy_id) contributing to each route group; (2) re-estimated the association with a GEE logistic model clustered by patient; (3) restricted this model to the narrower NPO-only-active subset, since tube-feeding and parenteral-nutrition patients may already have an alternate access device in place; (4) cross-tabulated formulation string against route classification, which showed every controlled-release (CR) dose in the altered-access subset was ordered oral-only; and (5) re-estimated the clustered model restricted to immediate-release (non-CR) formulations only, to remove this formulation confound. A final sensitivity excluded the single largest oral-only pharmacy order (24 of 71 oral-only doses) to confirm the clustered estimate was not attributable to one dominant order. eTable 17 reports the full ladder; the formulation-by-route cross-tabulation and a structured audit of the 71 oral-only doses (admission, pharmacy order, and formulation string) are also included.

#### S1.8 Statistical Model Specification

The ICU-versus-ward comparison (main-text) was fit as a GEE logistic regression with no covariates beyond ICU exposure (binomial family, logit link, exchangeable working correlation, clustered by subject_id):

mistimed ~ icu_exposed

fit on all 39,322 era-restricted doses, reported as a patient-clustered odds ratio, not an adjusted odds ratio, since the model contains no other covariates (eTable 12b for the primary delayed-or-omitted outcome; eTable 10b for the symmetric mistimed outcome). The within-admission comparison restricted to the 249 admissions with doses in both settings and compared each admission's ICU delayed-or-omitted rate to its own ward rate with a two-sided Wilcoxon signed-rank test.

The access-type categorical model (main-text primary) progressed through 3 specifications, each clustered by subject_id on the same 6,201 ICU-exposed doses, with normal/texture-modified access as the reference level: unadjusted (mistimed ~ C(access_level, Treatment(reference='normal'))); era-adjusted (adding C(era)); and fully adjusted (the version reported in the main text, Figure 3, and Results):

mistimed ~ C(access_level, Treatment(reference='normal')) + vent_flag_int + sedative_flag_int + C(era)

so that the access-type, ventilation, and sedative-infusion coefficients reported together in Figure 3 come from one specification rather than 2 separately-fit models, a distinction a round-4 reviewer critique specifically raised (eTable 13 reports all 3 specifications, the joint Wald test on the fully adjusted model's access-category coefficients, and the fully adjusted model's model-based marginal probabilities by access category, eTable 9). The pooled altered-access model (secondary, matching round 2/3 of this study) was:

mistimed ~ altered_access_int + vent_flag_int + sedative_flag_int + C(era)

fit on the same 6,201 doses (eTable 15 reports the full model including era terms omitted from the main-text figures for clarity); this model contains multiple covariates and is reported as an adjusted odds ratio (aOR). The same pooled formula was re-fit substituting the narrow NPO-only flag for altered_access_int (eTable 14). The ventilation term was also fit with and without altered_access_int in the model, using the reduced formula mistimed ~ vent_flag_int and mistimed ~ vent_flag_int + altered_access_int, to characterize a mediation-consistent pattern (eTable 15).

The comparator-medication interaction model is described in S1.6 (eTable 16). The route-order association was tested with the GEE models described in S1.7 (eTable 17) and, for the naive comparison only, a chi-square test, Cramer V, and a Wald 95% confidence interval computed directly from the 2x2 cell counts.

Approximate E-values (VanderWeele and Ding, *Ann Intern Med* 2017) were computed for the headline odds ratios, converting each odds ratio to a risk-ratio scale via the square-root approximation (appropriate because the mistimed outcome is not rare, at approximately 20% overall) and applying the standard E-value formula to both the point estimate and the confidence-interval bound closer to the null (eTable 11).

All GEE models used statsmodels 0.14 in Python 3.9; coefficients were exponentiated to odds ratios and 95% confidence intervals were exponentiated from the model's Wald confidence intervals on the log-odds scale.

#### S1.9 Delay-Threshold and Symmetric-Timing Sensitivity Analysis

The primary dosing-fidelity classification used a 60-minute delay threshold for "Administered"-coded records lacking an explicit delay flag. As a sensitivity analysis, the same classification was repeated at 15-, 30-, and 90-minute thresholds (eTable 12); the 15-minute threshold matches contemporary Parkinson's Foundation hospital-care guidance. The ICU-versus-ward unadjusted comparison was repeated at all four thresholds.

Because doses administered substantially early were rolled into the on-time category throughout, a symmetric "mistimed" definition (delayed, omitted, or early beyond the same window) was constructed at the 60- and 15-minute windows, and the ICU-versus-ward GEE comparison was re-estimated under each (eTable 10b). The distribution of early-administration magnitude, overall and by setting, is reported in eTable 10a.

#### S1.10 Route Cross-Validation Methodology

Because this pipeline has no access to clinical notes and no clinician available to adjudicate the true clinical appropriateness of an individual held or delayed dose, a structured-data substitute was used: for a random sample of doses with a classifiable ordered route, the emar_detail.route field (the per-dose administered route, distinct from prescriptions.route, the ordered route) was extracted and compared against the ordered-route classification. This checks internal consistency between two independently recorded structured signals; it does not establish clinical appropriateness. Of 1,833 sampled route-flexible doses, 1,813 (98.9%) had a documented emar_detail.route value, of which 193 (10.6%) recorded a non-oral administered route. Of 167 sampled oral-only doses, none had a documented emar_detail.route value, consistent with a nursing-documentation convention of recording route only when it deviates from a default oral swallow; this pattern meant a matched oral-only comparison arm could not be constructed for this cross-validation (eTable 18).

#### S1.11 Landmark Outcome and Ward-to-ICU Transfer Design Detail

To reduce reverse-causation from measuring dosing-fidelity burden and length of stay or mortality over the same, variable-length admission window, the whole-admission tertile design used in round 1 of this study was replaced with a 24-hour landmark design. Dosing-fidelity burden was computed only from doses scheduled in the first 24 hours of the admission (scheduletime ≤ admittime + 24h). Admissions with dischtime ≤ admittime + 24h (that is, admissions that died or were discharged within the landmark window) were excluded entirely from the landmark cohort, leaving 1,449 of the 1,665 era-restricted admissions. Length of stay was measured from the 24-hour landmark forward, not from admission. The coarse ICD-9/10-coded outcome composite (aspiration pneumonitis, delirium, neuroleptic malignant syndrome or malignant hyperthermia; codes in S3) was joined at the whole-admission level, because diagnoses_icd in MIMIC-IV carries no timestamp or present-on-admission flag; this composite is therefore not restricted to events occurring after the landmark and is reported throughout as a whole-admission, non-temporally-resolved association, never as a temporally sequenced outcome. Results are reported in eTable 7.

The same landmark cohort was reused, with exposure and outcome reversed, to test whether landmark-window dosing-fidelity burden predicted a subsequent, previously unobserved ICU transfer. Admissions were eligible if they survived to the 24-hour landmark and had not yet had any ICU stay begin by the landmark (icustays.intime ≤ admittime + 24h excluded the admission); 1,302 of 1,665 admissions met this ward-only-through-landmark criterion, of which 1,226 had ≥1 scheduled dose in the landmark window and so contributed a burden estimate. For each of two outcome windows (24 and 48 hours after the landmark), an admission was classified as having the outcome if its first ICU transfer (icustays.intime) fell within the window; an admission was included in that window's analytic set only if it either had the outcome or remained hospitalized (dischtime after the window closed) through the full window, so that early alive discharge without a transfer was not miscounted as a confirmed non-event. A GEE logistic model (icu_transfer ~ any_landmark_mistimed_dose, clustered by subject_id) was fit separately at each window. This design removes reverse causation from the timing of measurement (the exposure window strictly precedes the outcome window) but cannot exclude confounding by an unmeasured deterioration trajectory that would independently predict both a missed dose and a later transfer; its result (eTable 8) is reported as exploratory and hypothesis-generating only.

#### S1.12 Discharge-Summary Text Concordance for the Access-Type Variable

The access-type variable (S1.5) is derived from a single structured field (chartevents itemid 224001) and, as noted there, records what was charted rather than a clinician-adjudicated determination. This study has no clinician available to perform chart review, so a text-based, automated substitute was used: for each of the 290 ICU-exposed admissions in the access-type cohort, the dominant (modal) per-dose access category was computed using the identical 12-hour lookback classification the primary model fits on (S1.5, S1.8; ties broken toward the more-restricted category), then compared against that admission's discharge summary in MIMIC-IV-Note (deidentified free-text clinical notes, version 2.2). MIMIC-IV-Note ships only discharge summaries and radiology reports, not nursing or physician progress notes, so this check is admission-level and retrospective, describing whether the discharge narrative corroborates the dominant category anywhere in the hospital-course text, not whether every individual dose was correctly classified. It is a text-concordance check, not a chart review, and should not be read as clinician adjudication. Category-specific keyword patterns (case-insensitive) were: NPO, "nothing by mouth," or "nil per os" for the NPO category; "tube feeding," PEG, NGT, "nasogastric tube," gastrostomy, jejunostomy, or "enteral feeding/nutrition" for tube feeding; TPN, PPN, or "parenteral nutrition" for TPN/PPN. For the normal/texture-modified category, the check was reversed: the proportion of admissions with no restriction-related keyword anywhere in the discharge summary (a specificity check, since no restriction was coded). A discharge summary was available for 160 of 290 admissions (55.2%); admissions without one were excluded from the corresponding row. Results are reported in eTable 20.

### S2 Supplementary Tables

**eTable 1. Era-completeness comparison and 2014-2022 sensitivity.**

*1a. Included versus excluded-no-EMAR admissions, by calendar era.* Admissions meeting the primary diagnosis-plus-order definition (before era restriction), compared by whether they had a scheduled EMAR entry for levodopa. This comparison motivated restricting the analytic cohort to the 2011-2022 anchor-year-group eras.

| Characteristic | Included (EMAR present), n = 2,145 | Excluded (no EMAR), n = 1,444 |
| --- | --- | --- |
| Age, median, y | 74.0 | 76.5 |
| Male, % | 60.1 | 59.6 |
| In-hospital mortality, % | 3.1 | 2.8 |
| Era 2008-2010, % | 22.4 | 69.0 |
| Era 2011-2013, % | 17.2 | 27.4 |
| Era 2014-2016, % | 26.8 | 2.6 |
| Era 2017-2019, % | 20.0 | 0.6 |
| Era 2020-2022, % | 13.6 | 0.3 |

*1b. Sensitivity restricting the primary cohort to 2014-2022 (dropping the smaller residual imbalance in 2011-2013).*

| Cohort | n doses | n admissions | On time, % | Delayed, % | Omitted, % |
| --- | --- | --- | --- | --- | --- |
| Primary (2011-2022) | 39,322 | 1,665 | 79.8 | 13.9 | 6.4 |
| Restricted (2014-2022) | 31,711 | 1,295 | 80.1 | 13.5 | 6.4 |

| Cohort | ICU delayed-or-omitted, % | Ward delayed-or-omitted, % |
| --- | --- | --- |
| Primary (2011-2022) | 16.1 | 21.0 |
| Restricted (2014-2022) | 15.7 | 20.9 |

**eTable 2. STROBE item map.** Each STROBE checklist item for observational cohort studies is mapped to its location in the manuscript.

| STROBE item | Manuscript location |
| --- | --- |
| Title and abstract | Title page; Abstract |
| Background/rationale | Introduction, paragraphs 1-2 |
| Objectives | Introduction, paragraph 3 |
| Study design | Methods, Study Design and Setting |
| Setting | Methods, Study Design and Setting |
| Participants | Methods, Cohort Identification |
| Variables | Methods, The Dosing-Fidelity Measure; Access-Type Classification |
| Data sources/measurement | Methods, The Dosing-Fidelity Measure; eMethods S1.3-S1.7 |
| Bias | Discussion, Limitations |
| Study size | Results, Cohort |
| Quantitative variables | Methods, Access-Type Classification; Landmark Outcomes |
| Statistical methods | Methods, Statistical Analysis; eMethods S1.8-S1.9 |
| Participants (flow) | Results, Cohort; Figure 1 |
| Descriptive data | Results, Cohort; Table 1 |
| Outcome data | Results, Dosing Fidelity; Landmark Outcomes |
| Main results | Results, all subsections |
| Other analyses | Results, Sensitivity Analyses; eTables 5, 11-13, 16 |
| Key results | Discussion, paragraph 1 |
| Limitations | Discussion, Limitations |
| Interpretation | Discussion, paragraphs 2-4 |
| Generalisability | Discussion, Limitations |
| Funding | Title page |

**eTable 3. Levodopa formulary strings matched in prescriptions.drug.** Case-insensitive substring match on "carbidopa"; formulations captured are listed for reference. Every distinct matched string, including the generic-labeled row, was individually reviewed against its formulary description and confirmed to be a genuine carbidopa-levodopa combination product (S1.2).

Formulation string (as recorded in MIMIC-IV)

Carbidopa-Levodopa (25-100)

Carbidopa-Levodopa (25-250)

Carbidopa-Levodopa (10-100)

Carbidopa-Levodopa CR (25-100)

Carbidopa-Levodopa CR (50-200)

Carbidopa-Levodopa (25-100) ODT

carbidopa (generic entries)

**eTable 4. Fine-grained dose-classification breakdown, era-restricted cohort (N = 39,322 doses).** The primary three-way category (on time, delayed, omitted) used in the main text rolls up "early" and "missing timestamp" into on time, and "not given" and "hold dose" into omitted.

| Detail category | No. | % | Rolls up to (primary category) |
| --- | --- | --- | --- |
| On time | 20,103 | 51.1 | On time |
| Early (negative delta) | 11,263 | 28.6 | On time |
| Missing timestamp, coded Administered | 0 | 0.0 | On time |
| Delayed (≥60 min) | 5,449 | 13.9 | Delayed |
| Not Given | 2,412 | 6.1 | Omitted |
| Hold Dose | 95 | 0.2 | Omitted |
| Partial Administered | 0 | 0.0 | (none observed) |

**eTable 5. NPO-only lookback-window sensitivity and diet-documentation coverage, among ICU-exposed doses (S1.5).**

*5a. Association at each lookback window.* Documentation coverage for each window is reported in eTable 5b.

| Lookback | n doses NPO-flagged | Odds ratio | 95% CI | *P* value |
| --- | --- | --- | --- | --- |
| 6 h | 1,548 | 2.64 | 1.998-3.477 | <.001 |
| 12 h (primary) | 1,807 | 2.65 | 1.993-3.526 | <.001 |
| 24 h | 1,830 | 2.67 | 2.004-3.556 | <.001 |

*5b. Full documentation-coverage breakdown at each lookback window (% of the 6,201 ICU-exposed doses).*

| Lookback | NPO documented | Other diet-type documented | No recent documentation |
| --- | --- | --- | --- |
| 6 h | 25.0 | 55.2 | 19.9 |
| 12 h (primary) | 29.1 | 67.7 | 3.2 |
| 24 h | 29.5 | 69.3 | 1.2 |

**eTable 6. Chartevents diet-type documentation coverage, by whether the admission had any ICU stay (S1.5).** Diet-type documentation (chartevents itemid 224001) is charted by the ICU nursing system; this table quantifies the resulting coverage gap that limits the comparator-medication specificity analysis to ICU-exposed doses (S1.6).

| Admission group | n admissions | With ≥1 diet-type entry, No. (%) | With no diet-type entry, No. (%) |
| --- | --- | --- | --- |
| ICU-exposed (≥1 ICU stay) | 325 | 322 (99.1) | 3 (0.9) |
| No ICU stay | 1,340 | 0 (0.0) | 1,340 (100.0) |

No admission without an ICU stay contributed a single chartevents diet-type entry. Absence of an entry was classified with normal access by construction (S1.5); for ICU-exposed admissions this is a reasonable default (99.1% have direct documentation to draw on), but for admissions with no ICU stay, "normal access" reflects the complete absence of a source table rather than a documented normal diet. This is why the comparator-medication analysis (eTable 16) restricts its valid specificity test to ICU-exposed doses, and reports the full-cohort (ICU-plus-ward) version as descriptive only.

**eTable 7. Landmark outcomes by dosing-fidelity burden tertile (24-hour landmark; N = 1,449 admissions).**

| Tertile | n admissions | LOS from landmark, median (IQR), d |
| --- | --- | --- |
| Low burden | 708 | 3.2 (1.6-6.2) |
| Mid burden | 399 | 3.2 (1.5-6.7) |
| High burden | 342 | 4.1 (2.1-7.7) |

| Tertile | In-hospital mortality, No. (%) | ICD-coded composite, No. (%) |
| --- | --- | --- |
| Low burden | 14 (2.0) | 84 (11.9) |
| Mid burden | 13 (3.3) | 56 (14.0) |
| High burden | 11 (3.2) | 57 (16.7) |

LOS: epsilon-squared 0.007, BH-adjusted *P* = .006. Mortality: Cramer V 0.039, BH-adjusted *P* = .32. ICD-coded composite: Cramer V 0.056, BH-adjusted *P* = .15.

**eTable 8. Ward-to-ICU proximal-transfer analysis (24-hour landmark, exposure and outcome reversed; S1.11).** Exploratory and hypothesis-generating; cannot exclude confounding by an unmeasured deterioration trajectory.

*8a. Followable admissions and events at each outcome window.*

| Outcome window | n followable admissions | n new ICU transfers | % |
| --- | --- | --- | --- |
| 24 h | 993 | 12 | 1.2 |
| 48 h | 812 | 20 | 2.5 |

*8b. ICU transfer rate by landmark-window dosing-fidelity exposure.*

| Outcome window | Exposure group | n admissions | ICU transfer, % |
| --- | --- | --- | --- |
| 24 h | ≥1 mistimed dose in landmark window | 518 | 1.4 |
| 24 h | All doses on time in landmark window | 475 | 1.1 |
| 48 h | ≥1 mistimed dose in landmark window | 425 | 2.4 |
| 48 h | All doses on time in landmark window | 387 | 2.6 |

*8c. GEE model (clustered by patient).*

| Outcome window | Odds ratio | 95% CI | *P* value |
| --- | --- | --- | --- |
| 24 h | 1.29 | 0.406-4.079 | .667 |
| 48 h | 0.91 | 0.376-2.196 | .831 |

**eTable 9. Model-based predicted probability of a delayed-or-omitted dose, by access category (S1.8).** Predicted from the fully adjusted, patient-clustered GEE model (access category, mechanical ventilation, sedative infusion, era), holding ventilation, sedative infusion, and era at their observed sample means. Underlies Figure 4.

| Access category | Predicted probability, delayed or omitted, % |
| --- | --- |
| Normal or texture-modified diet | 15.3 |
| Tube feeding | 10.1 |
| TPN or PPN | 20.6 |
| NPO | 25.4 |

**eTable 10. Early-administration distribution and symmetric-timing sensitivity.**

*10a. Early-administration magnitude, overall and by setting.*

| Group | n doses | n early, % | Median early, min (IQR) |
| --- | --- | --- | --- |
| All | 39,322 | 28.6 | 17 (8-32) |
| ICU-exposed | 6,201 | 32.4 | 19 |
| Ward | 33,121 | 27.9 | 17 |

| Group | % ≥15 min early | % ≥30 min early | % ≥60 min early | % ≥90 min early |
| --- | --- | --- | --- | --- |
| All | 16.3 | 8.0 | 0.3 | 0.02 |
| ICU-exposed | 19.4 | 9.8 | 0.3 | 0.02 |
| Ward | 15.7 | 7.7 | 0.3 | 0.02 |

*10b. ICU-versus-ward comparison under a symmetric mistimed definition (delayed, omitted, or early beyond the same window), patient-clustered GEE.*

| Window | ICU mistimed, % | Ward mistimed, % | Odds ratio | 95% CI | *P* value |
| --- | --- | --- | --- | --- | --- |
| 60 min | 16.3 | 21.3 | 0.86 | 0.74-1.01 | .063 |
| 15 min | 68.1 | 69.0 | 1.05 | 0.95-1.15 | .328 |

**eTable 11. E-values for headline odds ratios (S1.8).** The E-value is the minimum risk-ratio-scale strength an unmeasured confounder would need with both the exposure and the outcome to fully explain away the point estimate, or to move the stated confidence-interval bound to the null.

| Estimate | Odds ratio | 95% CI | CI bound scored |
| --- | --- | --- | --- |
| ICU exposure (patient-clustered) | 0.87 | 0.74-1.01 | Upper |
| NPO (fully adjusted) | 1.89 | 1.36-2.62 | Lower |
| Tube feeding (fully adjusted) | 0.62 | 0.42-0.92 | Upper |
| Altered access, widened (adjusted) | 1.43 | 1.09-1.87 | Lower |
| Altered access, NPO-only (unadjusted) | 2.24 | 1.88-2.67 | Lower |
| Mechanical ventilation (adjusted) | 0.71 | 0.56-0.90 | Upper |
| Route order, naive (unadjusted) | 2.72 | 1.54-4.78 | Lower |
| Route order, clustered | 2.33 | 1.27-4.26 | Lower |

| Estimate | E-value, point estimate | E-value, CI bound |
| --- | --- | --- |
| ICU exposure (patient-clustered) | 1.35 | 1.08 |
| NPO (fully adjusted) | 2.09 | 1.61 |
| Tube feeding (fully adjusted) | 1.84 | 1.25 |
| Altered access, widened (adjusted) | 1.68 | 1.26 |
| Altered access, NPO-only (unadjusted) | 2.36 | 2.08 |
| Mechanical ventilation (adjusted) | 1.66 | 1.29 |
| Route order, naive (unadjusted) | 2.68 | 1.79 |
| Route order, clustered | 2.42 | 1.51 |

**eTable 12. Delay-threshold sensitivity analysis, era-restricted cohort.** The omitted proportion does not vary by threshold because omission is coded directly in the EMAR event field.

*12a. Dose-level fidelity proportions, No. (%).*

| Threshold | On time | Delayed | Omitted |
| --- | --- | --- | --- |
| 15 min | 18,678 (47.5) | 18,137 (46.1) | 2,507 (6.4) |
| 30 min | 23,925 (60.8) | 12,890 (32.8) | 2,507 (6.4) |
| 60 min (primary) | 31,366 (79.8) | 5,449 (13.9) | 2,507 (6.4) |
| 90 min | 34,648 (88.1) | 2,167 (5.5) | 2,507 (6.4) |

*12b. Delayed-or-omitted rate by setting, %.*

| Threshold | ICU-exposed | Ward |
| --- | --- | --- |
| 15 min | 48.6 | 53.2 |
| 30 min | 34.3 | 40.1 |
| 60 min (primary) | 16.1 | 21.0 |
| 90 min | 10.1 | 12.2 |

**eTable 13. Access-type categorical model, among ICU-exposed doses (S1.5, S1.8).** Normal or texture-modified access is the reference category. TPN/PPN estimates (35 doses, 2 patients) are shown for completeness but are not interpreted inferentially (Limitations).

*13a. Descriptive proportions.*

| Access type | n doses | n patients | Mistimed, % |
| --- | --- | --- | --- |
| Normal or texture-modified | 1,275 | 152 | 17.0 |
| Tube feeding | 3,084 | 73 | 9.6 |
| TPN/PPN | 35 | 2 | 20.0 |
| NPO | 1,807 | 184 | 26.3 |

*13b. GEE model, unadjusted, era-adjusted, and fully adjusted, versus normal access.* The fully adjusted model (access category, mechanical ventilation, sedative infusion, era, all in one prespecified specification) is the model cited in the main text (Figure 3; Results).

| Model | Term | Odds ratio | 95% CI | *P* value |
| --- | --- | --- | --- | --- |
| Unadjusted | Tube feeding | 0.52 | 0.341-0.792 | .002 |
| Unadjusted | TPN/PPN | 1.22 | 0.889-1.671 | .219 |
| Unadjusted | NPO | 1.74 | 1.243-2.431 | .001 |
| Era-adjusted | Tube feeding | 0.56 | 0.395-0.802 | .001 |
| Era-adjusted | TPN/PPN | 1.31 | 0.886-1.945 | .175 |
| Era-adjusted | NPO | 1.79 | 1.301-2.469 | <.001 |
| Fully adjusted | Tube feeding | 0.62 | 0.424-0.921 | .018 |
| Fully adjusted | TPN/PPN | 1.44 | 0.898-2.308 | .130 |
| Fully adjusted | NPO | 1.89 | 1.363-2.624 | <.001 |
| Fully adjusted | Mechanical ventilation | 0.74 | 0.512-1.086 | .126 |
| Fully adjusted | Active sedative infusion | 1.03 | 0.669-1.582 | .899 |

*13c. Joint Wald test on the 3 access-category coefficients' cluster-robust covariance (fully adjusted model).* Replaces a round-4 omnibus Kruskal-Wallis test that had been run on raw dose-level values without accounting for patient clustering.

| Test | Statistic | df | *P* value |
| --- | --- | --- | --- |
| Joint Wald, access category | χ² = 72.47 | 3 | <.001 |

**eTable 14. Altered oral access: narrow (NPO-only) versus widened definition, among ICU-exposed doses.** Both definitions fit in the same adjusted model (altered access, ventilation, sedative infusion, calendar era; clustered by patient), on the same era-restricted dose set. This pooled binary definition is a secondary comparison; the categorical model (eTable 13) is primary (S1.5).

| Definition | n doses flagged active | Odds ratio | 95% CI | *P* value |
| --- | --- | --- | --- | --- |
| Narrow (NPO substring only) | 1,807 | 2.24 | 1.88-2.67 | <.001 |
| Widened (NPO, tube feeding, TPN, or PPN) | 4,926 | 1.43 | 1.09-1.87 | .009 |

**eTable 15. Full adjusted models (pooled altered-access definition), S1.8.** All models are generalized estimating equations logistic regressions clustered by patient (binomial family, exchangeable working correlation).

*15a. ICU exposure and delayed-or-omitted dosing (all era-restricted doses, N = 39,322).* Covariate-free model; reported as a patient-clustered, not adjusted, odds ratio.

| Term | Odds ratio | 95% CI | *P* value |
| --- | --- | --- | --- |
| ICU-exposed (ref: ward) | 0.87 | 0.74-1.01 | .075 |

*15b. Primary pooled-access adjusted model among ICU-exposed doses (N = 6,201 doses, 290 admissions), with era terms.*

| Term | Odds ratio | 95% CI | *P* value |
| --- | --- | --- | --- |
| Altered oral access (widened) | 1.43 | 1.09-1.87 | .009 |
| Mechanical ventilation | 0.71 | 0.56-0.90 | .004 |
| Active sedative infusion | 1.11 | 0.82-1.50 | .496 |
| Admission era, 2014-2016 (ref: 2011-2013) | 0.89 | 0.55-1.44 | .630 |
| Admission era, 2017-2019 | 0.87 | 0.51-1.48 | .605 |
| Admission era, 2020-2022 | 0.95 | 0.56-1.62 | .857 |

*15c. Mechanical ventilation, with and without altered oral access in the model.*

| Model | Odds ratio | 95% CI | *P* value |
| --- | --- | --- | --- |
| Ventilation alone | 0.76 | 0.63-0.92 | .004 |
| Ventilation + altered oral access | 0.73 | 0.60-0.90 | .003 |

*15d. Unadjusted association between ventilation and altered oral access, among ICU-exposed doses.* Cramer V = 0.34, chi-square *P* < .001. Altered-access rate was 98.6% among ventilated doses versus 69.6% among non-ventilated doses.

**eTable 16. Comparator medication (statin) specificity analysis, S1.6.** Only the ICU-restricted rows are a valid specificity test; the "All doses" (ICU-plus-ward) rows are descriptive only, because the chartevents diet-type source cannot flag a ward-only admission as altered access regardless of true diet status (eTable 6).

*16a. Dose counts and mistimed rate, shared cohort (653 admissions, 394 patients).*

| Medication | Access status | n doses | n patients | Mistimed, % |
| --- | --- | --- | --- | --- |
| Levodopa | Normal access | 14,094 | 391 | 21.4 |
| Levodopa | Altered access | 3,003 | 81 | 14.7 |
| Statin (comparator) | Normal access | 3,367 | 389 | 24.5 |
| Statin (comparator) | Altered access | 529 | 66 | 11.0 |

*16b. GEE interaction models (medication x altered access, clustered by patient).* Strata: "ICU, unadj." = ICU-exposed only, unadjusted, N = 4,282 (valid); "ICU, vent.-adj." = ICU-exposed only, ventilation-adjusted, N = 4,282 (valid); "All doses" = ICU and ward, N = 20,993 (descriptive only, not a specificity test).

| Stratum | Term | Odds ratio | 95% CI | *P* value |
| --- | --- | --- | --- | --- |
| ICU, unadj. | Medication x access | 1.27 | 0.748-2.162 | .375 |
| ICU, vent.-adj. | Medication x access | 1.28 | 0.754-2.178 | .359 |
| All doses (descriptive) | Medication (levodopa vs statin) | 0.84 | 0.743-0.956 | .008 |
| All doses (descriptive) | Altered access (statin) | 0.38 | 0.219-0.658 | .001 |
| All doses (descriptive) | Medication x access | 1.66 | 1.214-2.267 | .002 |

**eTable 17. Route-order evidence ladder and formulation confound.**

*17a. Route-order association, oral-only vs route-flexible, among ICU-exposed doses with altered oral access active.* All rows are the same underlying comparison estimated under progressively stricter conditions; see S1.7.

| Analysis | n doses | Odds ratio | 95% CI | *P* value |
| --- | --- | --- | --- | --- |
| Naive (unadjusted, unclustered 2x2) | 4,601 | 2.72 | 1.54-4.78 | <.001 |
| Clustered, all doses | 4,601 | 2.33 | 1.27-4.26 | .006 |
| Clustered, NPO-only subset | 1,528 | 2.70 | 1.48-4.90 | .001 |
| Clustered, excl. largest order | 4,577 | 2.79 | 1.45-5.37 | .002 |
| Clustered, IR formulations only | 4,562 | 0.72 | 0.30-1.73 | .465 |

Full model names: "Naive" is an unadjusted, unclustered dose-level 2x2 odds ratio; "Clustered" rows are patient-clustered GEE models; "IR" is immediate-release.

| Analysis | n oral-only doses | Oral-only patients / orders |
| --- | --- | --- |
| Naive | 71 | 20 / 26 |
| Clustered, all doses | 71 | 20 / 26 |
| Clustered, NPO-only subset | 49 | 19 / 25 |
| Clustered, excl. largest order | 47 | 19 / 25 |
| Clustered, IR formulations only | 32 | 4 / 5 |

| Analysis | n route-flexible doses | Route-flexible patients / orders |
| --- | --- | --- |
| Naive | 4,530 | 173 / 364 |
| Clustered, all doses | 4,530 | 173 / 364 |
| Clustered, NPO-only subset | 1,479 | 169 / 308 |
| Clustered, excl. largest order | 4,530 | 173 / 364 |
| Clustered, IR formulations only | 4,530 | 173 / 364 |

*17b. Formulation-by-route cross-tabulation, altered-access-active ICU doses.* Every controlled-release (CR) dose was ordered oral-only; CR tablets cannot be crushed for enteral administration without destroying their extended-release mechanism.

| Formulation | Route-flexible, No. | Oral-only, No. |
| --- | --- | --- |
| Carbidopa-Levodopa (25-100) | 3,716 | 32 |
| Carbidopa-Levodopa (25-100) ODT | 576 | 0 |
| Carbidopa-Levodopa (25-250) | 104 | 0 |
| Carbidopa-Levodopa (25-250) ODT | 105 | 0 |
| Carbidopa-Levodopa (10-100) | 29 | 0 |
| Carbidopa-Levodopa CR (25-100) | 0 | 30 |
| Carbidopa-Levodopa CR (50-200) | 0 | 9 |

*17c. Structured audit of the 71 oral-only doses, by admission, pharmacy order, and formulation.* The closest MIMIC-only substitute for a manual chart audit, since this pipeline has no clinical-note or chart access. One admission (hadm_id 29401358, one pharmacy order) contributed 24 of the 71 doses (33.8%); eTable 17a's "excluding largest single order" row confirms the clustered estimate does not depend on this one order.

| Admission (hadm_id) | Pharmacy order (pharmacy_id) | Formulation | n doses |
| --- | --- | --- | --- |
| 20748538 | 50344460 | Carbidopa-Levodopa CR (25-100) | 3 |
| 20959548 | 58494910 | Carbidopa-Levodopa CR (25-100) | 3 |
| 21260565 | 63153599 | Carbidopa-Levodopa CR (50-200) | 1 |
| 21294192 | 94562272 | Carbidopa-Levodopa CR (25-100) | 1 |
| 22330779 | 51603335 | Carbidopa-Levodopa CR (25-100) | 1 |
| 23058233 | 80251075 | Carbidopa-Levodopa CR (25-100) | 3 |
| 23509367 | 84971692 | Carbidopa-Levodopa CR (50-200) | 4 |
| 24022759 | 74699064 | Carbidopa-Levodopa (25-100) | 1 |
| 24182688 | 87302662 | Carbidopa-Levodopa CR (25-100) | 5 |
| 24275282 | 67052 | Carbidopa-Levodopa CR (25-100) | 1 |
| 24849809 | 47059060 | Carbidopa-Levodopa CR (50-200) | 1 |
| 24849809 | 73920037 | Carbidopa-Levodopa CR (25-100) | 1 |
| 25483863 | 10203618 | Carbidopa-Levodopa CR (25-100) | 1 |
| 25483863 | 14027185 | Carbidopa-Levodopa CR (25-100) | 1 |
| 25483863 | 98774099 | Carbidopa-Levodopa CR (25-100) | 2 |
| 26495589 | 40435653 | Carbidopa-Levodopa CR (25-100) | 2 |
| 26562583 | 71323174 | Carbidopa-Levodopa CR (25-100) | 1 |
| 27401101 | 44252515 | Carbidopa-Levodopa CR (50-200) | 3 |
| 27431422 | 18351900 | Carbidopa-Levodopa (25-100) | 1 |
| 27519233 | 36128838 | Carbidopa-Levodopa CR (25-100) | 1 |
| 27519233 | 69594616 | Carbidopa-Levodopa CR (25-100) | 1 |
| 28663748 | 33197653 | Carbidopa-Levodopa (25-100) | 4 |
| 28663748 | 45367149 | Carbidopa-Levodopa CR (25-100) | 1 |
| 28663748 | 94073581 | Carbidopa-Levodopa (25-100) | 2 |
| 29401358 | 25637593 | Carbidopa-Levodopa (25-100) | 24 |
| 29902541 | 59536018 | Carbidopa-Levodopa CR (25-100) | 2 |

**eTable 18. emar_detail route cross-validation sample.** "Documented" is the n (%) of the sampled doses with a non-blank emar_detail.route value; "Non-oral" is the n (%), of those documented, whose administered route was not PO/Oral.

| Ordered-route classification | n sampled | Documented | Non-oral |
| --- | --- | --- | --- |
| Route-flexible | 1,833 | 1,813 (98.9%) | 193 (10.6%) |
| Oral-only | 167 | 0 (0.0%) | not applicable |

**eTable 19. PD ascertainment sensitivity: primary (single-code) versus stricter (repeated-diagnosis) cohort.** The stricter cohort requires the Parkinson disease diagnosis code on 2 or more separate admissions for the same patient.

| Cohort | n doses | On time, % | Delayed, % | Omitted, % |
| --- | --- | --- | --- | --- |
| Primary (era-restricted, single-code) | 39,322 | 79.8 | 13.9 | 6.4 |
| Stricter (≥2 admissions with diagnosis code) | 28,750 | 80.4 | 13.5 | 6.1 |

**eTable 20. Discharge-summary text concordance for the dominant access-type category, ICU-exposed admissions with an available discharge note (S1.12).** For normal/texture-modified, the reported value is specificity (no restriction-related keyword in the discharge summary). For tube feeding and NPO, the reported value is sensitivity (a category-specific keyword present in the discharge summary). No admission had a dominant TPN/PPN category among those with a discharge note.

| Dominant category (n admissions with a note) | n corroborated | % |
| --- | --- | --- |
| Normal/texture-modified (60) | 53 | 88.3 |
| Tube feeding (32) | 23 | 71.9 |
| NPO (68) | 12 | 17.6 |

### S3 Codebook

**Diagnosis codes.** Idiopathic Parkinson disease: ICD-9 332.0; ICD-10 G20. Secondary/drug-induced parkinsonism (excluded): ICD-10 G21.x; ICD-9 332.1.

**ICD-coded outcome composite.** Whole-admission, non-temporally-resolved codes used to construct the composite reported in eTable 7 (S1.11).

| Outcome | ICD-9 | ICD-10 |
| --- | --- | --- |
| Aspiration pneumonitis | 507.0 | J69.0 |
| Delirium | 293.0 | F05.x |
| Neuroleptic malignant syndrome or malignant hyperthermia | 333.92, 995.86 | G21.0 |

**MIMIC-IV itemids used.**

| Covariate | Source table | Itemid(s) |
| --- | --- | --- |
| Diet type / altered oral access / access type | chartevents | 224001 |
| Invasive mechanical ventilation | procedureevents | 225792 |
| Propofol infusion | inputevents | 222168 |
| Midazolam infusion | inputevents | 221668 |
| Dexmedetomidine infusion | inputevents | 225150, 229420 |
| Fentanyl infusion | inputevents | 221744, 225942, 225972 |

**Diet-type value to access-type ordinal level (S1.5).**

| Ordinal level | Category | chartevents diet-type value(s) |
| --- | --- | --- |
| 0 | Normal/texture-modified | Any of 16 values^a^, or no entry |
| 1 | Tube feeding | Tube Feeding |
| 2 | TPN/PPN | TPN, PPN |
| 3 | NPO | NPO |

^a^The 16 distinct level-0 values: House - Regular, 2 gm sodium heart healthy, Pureed, Nectar Thick, Diabetic, Clear liquid, Sips, Soft solid, Thin liquid, Renal, NAS/low cholesterol, Full liquid, Honey thick, Ground, Lactose free, No liquid.

**Route strings and classification.**

| prescriptions.route value | Classification |
| --- | --- |
| PO/NG | Route-flexible |
| NG | Route-flexible |
| G TUBE | Route-flexible |
| J TUBE | Route-flexible |
| SL | Route-flexible |
| PO | Oral-only |
| ORAL | Oral-only |
| (other or missing) | Unknown (excluded from route-order analysis) |

**EMAR event-code classification.**

| event_txt value | Detail category | Primary category |
| --- | --- | --- |
| Administered | On time / Delayed / Early, by delta | On time* |
| Administered in Other Location | Same as Administered | On time* |
| Partial Administered | Partial (none observed) | On time |
| Delayed Administered | Delayed | Delayed |
| Not Given | Omitted, not given | Omitted |
| Hold Dose | Omitted, hold | Omitted |
| (no timestamp, Administered-coded) | Missing timestamp (none observed) | On time |

*Delayed instead if delta ≥ the threshold in use.

### S4 Code and Data Availability

MIMIC-IV v3.1 is available to credentialed researchers through PhysioNet (<https://physionet.org/content/mimiciv/>) after completion of human-subjects training and execution of the data use agreement; it cannot be redistributed. Analysis code (cohort extraction, dose-level and admission-level dataset construction, route and access classification, the access-type categorical and lookback-sensitivity analysis, the comparator-medication specificity analysis, the route-order re-analysis and formulation-confound check, landmark and ward-to-ICU-transfer dataset construction, E-value computation, statistical analysis, and figure generation) is available at <https://github.com/Alon-Gorenshtein/Levodopa-Administration-Timing-During-Hospitalization>. The repository does not contain patient-level data, consistent with the MIMIC-IV data use agreement.
